# AI Chatbots as Emerging Tools in Youth Mental Health Help-Seeking: Insights from New Jersey Youth

**DOI:** 10.64898/2026.06.02.26354131

**Authors:** Roxanne Alvarado-Torres, Irakli Kakauridze, Erika Bonnevie

## Abstract

Youth in the United States are experiencing growing mental health challenges, yet many face barriers to accessing timely, affordable, and stigma-free support. At the same time, artificial intelligence (AI) chatbots have become widely available and are increasingly being used by young people as tools for information seeking, coping, and self-reflection. This brief report explores how youth are engaging with AI chatbots for mental health support, drawing from qualitative interviews conducted in New Jersey. Nine semi-structured interviews were completed with participants ages 19–22. Thematic analysis revealed five core themes: (1) generational change, peer communication, and humor as coping and normalization tools; (2) internal and external barriers to self-recognition and help-seeking; (3) AI chatbots as a safe and accessible first step; (4) AI chatbots as a tool for filling information gaps; and (5) limits of AI chatbots and the preference for human connection. These findings indicate that young people see AI chatbots as private, judgment-free starting points for exploring their emotions and seeking early support. However, they also recognize that these tools cannot replace human connection or professional care. For public health, this presents both challenges and opportunities in utilizing the accessibility of AI chatbots while ensuring ethical design, cultural responsiveness, and protections that safeguard youth privacy and equity.

## Introduction

Teens and young adults in the U.S. are facing increasing mental health challenges. In 2021, 21% of youth aged 3-17 were diagnosed with a mental health condition, marking a 40% rise from a decade earlier (Agency for Healthcare Research and Quality (US) 2022). Nearly 30% of high school students reported poor mental health, while 36% of young adults aged 18-25 experienced mental illness in 2022, with 12% facing serious mental illness (National Institute of Mental Health 2024). New Jersey mirrors these trends, with 23% of children ages 3-17 having a mental, emotional, developmental, or behavioral condition (Lowrie and Schwartzman 2023). The 2023 New Jersey Young Adult Survey reported that 46% of respondents aged 18-25 experienced a mental health problem, highlighting the growing youth mental health crisis and the need for accessible support. Anxiety-related emergency room visits and hospitalizations for New Jersey teens increased 36% and 54% respectively between 2019-2021 (Lowrie and Shwartzman 2023). As the demand for care outpaces available services, interest is growing in digital tools that can expand access and reduce barriers to youth mental health support.

Digital mental health interventions delivered through telehealth, mobile apps, and social media campaigns are increasingly common complements to traditional care (Hamilton, Siegel, and Carper 2022; Potts et al. 2025). During the COVID-19 pandemic, telehealth use among teens surged, with 45% accessing it for mental health treatment by 2022 (Breslau et al. 2025). This approach is especially beneficial for youth in rural or underserved areas facing provider shortages and access barriers (Haleem et al. 2021; Orsolini et al. 2021). Mobile apps can also provide discreet support, with over half of teens reporting usage (Common Sense 2024), though issues like privacy concerns and inconsistent engagement persist (Koh, Tng, and Hartanto 2022). Crisis support lines are additional resources for adolescents who prefer text-based support (Crisis Text Line 2023), while social media campaigns aim to reduce stigma around mental health (Collins et al. 2020; Tam et al. 2024).

Artificial Intelligence (AI) chatbots are emerging as tools in mental health care. They vary in complexity, from simple rule-based systems that follow scripted flows to advanced conversational AI tools that use large language models to simulate human-like dialogue and generate personalized responses (Allouch, Azaria, and Azoulay 2021; Finn n.d.; Gillis and Hashemi-Pour 2024; IBM 2023, 2024). In controlled research settings, early studies suggest that AI chatbots may assist users with mood tracking, cognitive behavioral strategies, and psychoeducation (Balcombe 2023). For example, tools like Woebot, Wysa, and Tess were purpose-built for mental health and evaluated in controlled research settings, typically as part of structured interventions where researchers guided user engagement (Beatty et al. 2022; Fitzpatrick, Darcy, and Vierhile 2017; Fulmer et al. 2018). Studies on these platforms suggested that Wysa and Woebot provided meaningful support across a range of concerns, including managing academic stress (De Nieva et al. 2020), reducing anxiety among university students (Fitzpatrick et al. 2017), supporting individuals with ADHD (Jang et al. 2021), and alleviating depressive symptoms among highly engaged users (Feng et al. 2025; Inkster, Sarda, and Subramanian 2018).

However, these tools were specifically designed for mental health use and deployed under researcher oversight. In contrast, young people today are increasingly turning to general-purpose conversational AI tools such as ChatGPT, DeepSeek, and Claude for companionship, emotional support, and even informal therapeutic advice (Andoh 2025; Johns Hopkins Medicine 2025). Conversational AI chatbots are increasingly embedded in youth digital ecosystems and are being used to manage stress and support emotional well-being. A recent report indicates that 72% of teens ages 13-17 have used AI tools, with 13% reporting daily use and 8% engaging multiple times per day (Caldwell and Fisher 2025). One-third of teens reported using AI tools for social interaction or relationships, 18% used them for conversation or social practice, 12% for emotional or mental health support, and 9% described their AI as a “friend” or “best friend” (Caldwell and Fisher 2025). Youth describe these tools as helpful for stress management, coping strategies, and reframing techniques (Newport Academy 2024).Their appeal lies in the privacy they offer, their 24/7 availability, and their accessibility for youth in resource-limited settings. However, several concerns have been raised, including the inability of AI chatbots to replicate the emotional depth of human relationships, the risk of algorithmic bias affecting marginalized youth, potential data privacy issues, and the absence of clear safeguards in many platforms (American Psychological Association 2025; Marshall 2025; Smith, Bradbury, and Karney 2025). These factors point to the potential applications and risks of AI chatbots in mental health, yet little is known about how young people actually engage with these tools in everyday contexts.

The Public Good Projects (PGP) partnered with Partners in Prevention to explore how youth in New Jersey discuss and access mental health resources. This study specifically examines young people’s perspectives on conversational AI chatbots as emerging tools within the mental health support landscape. The aim of this study is to provide insights into how AI chatbots influence youth help-seeking behaviors. These insights can help inform the development of youth-centered digital interventions and guide the responsible integration of AI into mental health support systems.

## Methods

Semi-structured interviews with youth were conducted between April 30 and June 5, 2025.

Participants were recruited using a snowball sampling method. Through Partners in Prevention, the Youth Tobacco Action Group, a statewide youth-led program that engages young people across New Jersey to educate, advocate, and build leadership skills, was asked to share information about the study’s goals and the purpose of the interviews. The partner was asked to distribute a sign-up link to young people who might be interested in participating. All interviews were conducted in English via Zoom and lasted 30-45 minutes. One researcher facilitated and analyzed all interviews. Prior to each interview, the researcher reviewed the consent form verbally with participants to ensure comprehension. Verbal consent was obtained for both participation and audio recording, in accordance with IRB-approved procedures. With permission, interviews were recorded and later transcribed verbatim. Participants received an incentive for their time and contributions.

Interviews examined how young people understand, discuss, and manage their mental health, as well as their experiences with coping strategies and accessing support. Participants were asked specific questions about the use of AI chatbots, such as ChatGPT and similar tools, as resources for mental health. Additionally, they were invited to describe how these tools were being utilized. Thematic analysis was used to analyze the qualitative data, incorporating both inductive and deductive coding approaches. Codes and themes were developed and refined iteratively until thematic saturation was achieved A second researcher was consulted to review and validate the emerging themes. Transcripts and interview coding were managed and analyzed using Dedoose version 10. The study was approved by Advarra Institutional Review Board (Pro00085339).

### Findings

Nine semi-structured interviews were conducted with participants between the ages of 19 and 22. While gender identity was not formally collected, the sample appeared to include seven female-presenting and four male-presenting individuals, based on the interviewer’s observation. Thematic analysis of the interview transcripts yielded five key themes related to how youth perceive and engage with mental health resources: (1) generational change, peer communication, and humor as tools for coping and normalization; (2) internal and external barriers to self-recognition and help-seeking; (3) AI chatbots as a perceived safe and accessible first step; (4) AI chatbots as a means of filling information gaps; and (5) limitations of AI chatbots and a continued preference for human connection.

#### Generational Change, Peer Communication, and Humor as Coping and Normalization Tool

Participants consistently described their generation as more open and self-aware when discussing mental health compared to older generations. Peer communication emerged as a preferred channel for disclosure, with peers viewed as more relatable, empathetic, and less judgmental than adults. Humor frequently served as a coping mechanism and conversational entry point, with participants describing the use of casual or exaggerated jokes to express distress. While humor helped normalize mental health discussions within peer groups, some participants acknowledged that it could obscure more serious underlying issues. In contrast, several youth expressed frustration with older generations, who were often perceived as dismissive of the mental health challenges faced by young people. This generational disconnect was cited as a barrier to seeking support from adults.

#### Internal and External Barriers to Self-Recognition and Help-Seeking

Despite increased awareness of mental health, participants described minimizing their own struggles. Several noted that youth often postpone seeking support until challenges become unmanageable, at which point intervention feels more difficult or inaccessible. Common emotional experiences, such as loneliness or insecurity, were frequently dismissed as normal or expected, reinforcing the perception that their concerns were not serious enough to warrant professional attention.

A combination of internal and external factors contributed to this minimization. Externally, participants reported that adults often dismissed their experiences, suggesting they were too young to understand stress or characterizing their concerns as a typical part of adolescence. Internally, youth described feelings of embarrassment, fear of overreacting, and concern about burdening others. At the same time, some participants associated help-seeking with personal weakness and preferred to manage their emotions independently.

> *“I feel like we still struggle with* [mental health], *because at least a lot of the people I know like to shoulder their mental health themselves and have this “I can handle it on my own” mentality. Because of that, they shy away from discussing it with friends and family*.*”*

#### AI Chatbots as a Perceived Safe and Accessible First Step

Participants reported using conversational AI chatbots, such as ChatGPT and similar tools, as an initial step in managing mental health concerns. These tools were often accessed to determine whether their emotional experiences were typical or to find coping strategies before seeking support from another person. A key aspect of their appeal was the perception of chatbots as safe, anonymous, and nonjudgmental. The ability to engage privately reduced fears of stigma and allowed users to express thoughts they might otherwise keep to themselves. This sense of psychological safety made chatbots feel less intimidating than traditional sources of support, particularly for sensitive or uncertain concerns. Several participants likened chatbots to therapists or counselors, noting that their conversational style felt more personal and engaging than static online resources. Although youth recognized that chatbots lack the empathy of a human, many described the perceived neutrality and lack of judgment as emotionally protective, particularly in situations where opening up to peers or adults felt risky. This neutrality functioned as an “emotional cover,” creating space for unfiltered reflection and emotional processing.

> *“I like asking ChatGPT because it almost feels like a person to talk to*. […] *I think it’s better than just Googling something and getting thousands of results. With ChatGPT, you get one entry that’s tailored to your exact question*.*”*

Beyond emotional safety, youth also emphasized the functional advantages of chatbots. They valued the immediacy, clarity, and adaptability of responses, especially in contrast to the volume and inconsistency of search engine results. Participants described using chatbots to generate tailored advice, request information in different formats or languages, and receive actionable steps during moments of stress. Some turned to these tools during acute emotional episodes and found the suggestions, such as journaling or going outside, helpful in de-escalating distress. While not seen as substitutes for human connection or clinical care, chatbots were widely viewed as low-barrier tools that could offer practical support when users did not know where to begin.

#### AI Chatbots as a Tool for Filling Information Gaps

Youth participants described using AI chatbots to address gaps in the availability or accessibility of mental health information. They emphasized the value of being able to tailor responses to their specific needs, such as requesting simpler explanations, breaking down complex content into concrete steps, or reframing information in more relatable terms. One participant shared that they used a chatbot to translate information into Spanish, noting that this feature was especially helpful when other resources did not reflect their linguistic needs. Across interviews, participants viewed this adaptability as a key strength of AI chatbots, particularly in situations where culturally or linguistically relevant materials were limited. For these youth, chatbots offered a practical way to reshape information so it made sense in their own context, filling a gap often left by traditional health systems and online resources.

> *“I can manipulate it however I want - expand on it, translate it into Spanish, explain it differently, or give me three actionable steps. You can break it apart in any way you want, and that’s why we like it. It’s like Google, but quicker and more flexible”*

#### Limits of AI Chatbots and Preference for Human Connection

While participants acknowledged the usefulness of AI chatbots, there was broad consensus that these tools should not replace professional mental health care. Several participants emphasized that chatbots were not appropriate for diagnosing mental health conditions and that formal support from a therapist or doctor remained essential. Others highlighted that, despite the utility of chatbots, they lacked the emotional depth and empathy that human interaction provides. Many youth expressed a preference for personal, face-to-face conversations when dealing with more serious concerns. Concerns about the accuracy of chatbot responses were also common. Participants noted that information generated by AI tools could be unreliable, particularly because it draws on internet-based content. Some youth described strategies for verifying responses, such as cross-checking with other sources. Additionally, several participants raised questions about privacy and ethics, expressing discomfort about how their personal information might be collected, stored, or used by these platforms.

> *“The more and more you engage with it, the more data it collects about you and the information you give it. It’s interesting to think about, but I personally would prefer just having a person to speak to*.*”*

## Discussion

This study examined how youth in New Jersey access and discuss mental health resources, with particular attention to their use of general-purpose technologies such as conversational AI chatbots for emotional support. Participants described these tools as accessible, private, and nonjudgmental entry points for checking symptoms, processing emotions, and identifying coping strategies. Their reflections suggest that youth are incorporating AI chatbots into a broader help-seeking landscape that values open conversations about mental health and supports greater autonomy in managing emotional challenges. At the same time, youth articulated clear limitations, including concerns about the lack of human empathy, inconsistent accuracy, and data privacy. They consistently positioned chatbots as supplementary tools useful for initial reflection or guidance but not as replacements for professional mental health care.

These findings contribute insights from youth into how general technologies are being adapted for mental health support. By focusing on young people’s voices, this study provides a grounded perspective on how emerging digital tools intersect with broader trends in mental health help-seeking. The results highlight the value of youth involvement in the design and development of AI systems, underscoring the need for culturally responsive and ethically designed tools that reflect young people’s lived experiences and evolving needs. As digital platforms continue to shape mental health pathways, incorporating youth perspectives will be essential to ensuring that these tools are developed and deployed in ways that prioritize safety, protect privacy, and minimize potential harm.

From a public health perspective, the challenge lies in balancing the accessibility of AI chatbots with the imperative for responsible design and deployment. Ethical frameworks, transparent development processes, and robust safeguards have been cited as essential to ensure that these tools improve the quality and safety of mental health support (Mennella et al. 2024). Others have also highlighted that structural inequities in the data used to train large language models deserve close scrutiny, as these models often mirror the biases, omissions, and power dynamics present in the broader digital information ecosystem (Pava et al. 2022). For example, mental health content online is disproportionately produced in high-income, English-speaking contexts, which can lead to underrepresentation of perspectives from historically marginalized communities, including racial and ethnic minorities, non-English speakers, and LGBTQ+ youth (Blease and Torous 2023; Lynch 2025). As a result, these tools may fail to capture culturally nuanced understandings of mental health or may reinforce dominant narratives that do not resonate with diverse users. Without deliberate efforts to diversify training data and critically assess model outputs, AI tools risk perpetuating the very inequities they aim to address.

AI chatbots may introduce significant potential harms, particularly in their interactions with young people. Recent reports and media coverage have raised growing concerns about how social and generative AI chatbots, which also function as companions, affect their users (Chatterjee 2025; Common Sense Media 2025; Hill 2025; Sanford 2025). These systems can blur the line between genuine and simulated relationships, fostering a misleading sense of emotional intimacy that may intensify feelings of loneliness, reinforce unhealthy coping mechanisms, or delay the pursuit of real-world help (Common Sense Media 2025).

Additionally, because AI chatbots are designed to please users, they may unintentionally validate harmful thoughts or fail to recognize language indicating a crisis, which leaves serious issues unaddressed(Common Sense Media 2025). Other reports highlight that the ease for users to bypass safety measures by using specific prompts. The longer users interact with the AI, the more adept they become at circumventing these safeguardss (Center for Countering Digital Hate 2025). Without proper protections in place, AI chatbots risk not only failing to support mental health but also exacerbating existing vulnerabilities.

Findings from this study reflect a potential shift in how youth approach help-seeking. Participants described using general-purpose conversational AI chatbots, not originally intended for clinical care, as tools to fill emotional and informational gaps. They viewed these platforms as distinct from traditional resources, offering low-barrier, judgment-free spaces where they could begin to articulate concerns and explore coping strategies in ways that felt safe and private. Future research should examine how youth from diverse backgrounds engage with AI chatbots, how identity and cultural context shape these perceptions, and how such tools might be integrated into broader mental health strategies that support, rather than replace, human connection and professional care.

## Limitations

This study has several limitations. The use of AI chatbots among youth was a secondary focus within a broader project on youth mental health in New Jersey, and as such, the research did not explore in depth why some young people engage with these tools, how frequently they use them, or in what contexts they find most beneficial. The sample size was small and non-representative, recruited through snowball sampling in partnership with a youth-focused organization. This recruitment method may have introduced selection bias, drawing participants already engaged with health-related topics and limiting the generalizability of findings. Additionally, the data relied on self-reported perceptions, which may be subject to social desirability bias or overestimation of digital literacy. Interviews were also conducted within a specific geographic and temporal context, which may not capture the perspectives of youth in other regions or as AI tools continue to evolve. Future research should involve larger and more diverse samples and explore how demographic factors such as age, gender, and cultural background influence youth engagement with AI chatbots.

## Conclusion

AI chatbots are becoming part of youth help-seeking pathways, valued for their accessibility, neutrality, and immediacy. They represent meaningful early steps in how youth search for information and manage mental health concerns. Future research with larger and more representative samples should investigate how youth across different demographics engage with AI tools, under what conditions they are most helpful, and how risks can be mitigated.

## Data Availability

Due to the sensitive nature of the qualitative interviews, supporting data are restricted to protect participant privacy and confidentiality.

## Notes

### Competing Interest Statement

The authors have declared no competing interest.

### Author Declarations

The study was approved by Advarra Institutional Review Board (Pro00085339).

## References

Agency for Healthcare Research and Quality (US). 2022. 2022 National Healthcare Quality and Disparities Report. Rockville, MD. https://www.ncbi.nlm.nih.gov/books/NBK587174/.

Allouch, Merav, Amos Azaria, and Rina Azoulay. 2021. “Conversational Agents: Goals, Technologies, Vision and Challenges.” Sensors (Basel, Switzerland) 21(24). doi:10.3390/s21248448.

American Psychological Association. 2025. “Artificial Intelligence and Adolescent Well-Being: An APA Health Advisory.” https://www.apa.org/topics/artificial-intelligence-machine-learning/health-advisory-ai-adolescent-well-being.

Andoh, Efua. 2025. “Many Teens Are Turning to AI Chatbots for Friendship and Emotional Support.” Monitor in Psychology, October 1.

Balcombe, Luke. 2023. “AI Chatbots in Digital Mental Health.” Informatics 10(4):82. doi:10.3390/informatics10040082.

Beatty, Clare, Tanya Malik, Saha Meheli, and Chaitali Sinha. 2022. “Evaluating the Therapeutic Alliance With a Free-Text CBT Conversational Agent (Wysa): A Mixed-Methods Study.” Frontiers in Digital Health 4. doi:10.3389/fdgth.2022.847991.

Blease, Charlotte, and John Torous. 2023. “ChatGPT and Mental Healthcare: Balancing Benefits with Risks of Harms.” BMJ Mental Health 26(1). doi:10.1136/bmjment-2023-300884.

Breslau, Joshua, Alyssa Burnett, Olesya Baker, Jonathan H. Cantor, Ryan K. McBain, Ateev Mehrotra, Jacquelin M. Rankine, Bradley D. Stein, Fang Zhang, and Hao Yu. 2025. “Telehealth Use for Mental Health Treatment Among US Adolescents.” Journal of Adolescent Health. doi:10.1016/j.jadohealth.2025.03.025.

Caldwell, Jennifer, and John H. N. Fisher. 2025. Talk, Trust, and Trade-Offs: How and Why Teens Use AI Companions. https://www.commonsensemedia.org/sites/default/files/research/report/talk-trust-and-trade-offs_2025_web.pdf.

Center for Countering Digital Hate. 2025. Fake Friend: How ChatGPT Betrays Vulnerable Teens by Encouraging Dangerous Behavior. https://counterhate.com/wp-content/uploads/2025/08/Fake-Friend_CCDH_FINAL-12Sep.pdf.

Chatterjee, Rhitu. 2025. “As More Teens Use AI Chatbots, Parents and Lawmakers Sound the Alarm about Dangers.” Mental Health, October 1.

Collins, R., N. Eberhart, R. Seelam, R. De Guttry, and M. Mizel. 2020. 2019 Evaluation of Los Angeles County’s WhyWeRise Mental Health Campaign. https://www.rand.org/pubs/research_reports/RR4441.html.

Common Sense. 2024. Getting Help Online: How Young People Find, Evaluate, and Use Mental Health Apps, Online Therapy, and Behavioral Health Information. https://www.commonsensemedia.org/sites/default/files/research/report/2024-getting-help-online-hopelab-report_final-release-for-web.pdf.

Common Sense Media. 2025. AI Risk Assessment: Social AI Companions. https://www.commonsensemedia.org/sites/default/files/pug/csm-ai-risk-assessment-social-ai-companions_final.pdf.

Crisis Text Line. 2023. A Decade Of Impact. https://www.crisistextline.org/wp-content/uploads/2023/10/A-Decade-of-Impact-Report.pdf.

Feng, Yi, Yaming Hang, Wenzhi Wu, Xiaohang Song, Xiyao Xiao, Fangbai Dong, and Zhihong Qiao. 2025. “Effectiveness of AI-Driven Conversational Agents in Improving Mental Health Among Young People: Systematic Review and Meta-Analysis.” Journal of Medical Internet Research 27(1). doi:10.2196/69639.

Finn, Teaganne. n.d. “6 Types of Chatbots and How to Choose the Right One for Your Business.”

Fitzpatrick, Kathleen Kara, Alison Darcy, and Molly Vierhile. 2017. “Delivering Cognitive Behavior Therapy to Young Adults With Symptoms of Depression and Anxiety Using a Fully Automated Conversational Agent (Woebot): A Randomized Controlled Trial.” JMIR Mental Health 4(2):e19. doi:10.2196/mental.7785.

Fulmer, Russell, Angela Joerin, Breanna Gentile, Lysanne Lakerink, and Michiel Rauws. 2018. “Using Psychological Artificial Intelligence (Tess) to Relieve Symptoms of Depression and Anxiety: Randomized Controlled Trial.” JMIR Mental Health 5(4):e64. doi:10.2196/mental.9782.

Gillis, Alexander S., and Cameron Hashemi-Pour. 2024. “What Is Conversational AI (Conversational Artificial Intelligence)?” https://www.techtarget.com/searchenterpriseai/definition/conversational-AI.

Haleem, Abid, Mohd Javaid, Ravi Pratap Singh, and Rajiv Suman. 2021. “Telemedicine for Healthcare: Capabilities, Features, Barriers, and Applications.” Sensors International 2:100117. doi:10.1016/j.sintl.2021.100117.

Hamilton, Jessica L., David M. Siegel, and Matthew M. Carper. 2022. “Digital Media Interventions for Adolescent Mental Health.” Pp. 389–416 in Handbook of Adolescent Digital Media Use and Mental Health. Cambridge University Press.

Hill, Kashmir. 2025. “A Teen Was Suicidal. ChatGPT Was the Friend He Confided In.” Technology, August 27.

IBM. 2023. “What Are Large Language Models (LLMs)?” https://www.ibm.com/think/topics/large-language-models. IBM. 2024. “What Is a Chatbot?”

Inkster, Becky, Shubhankar Sarda, and Vinod Subramanian. 2018. “An Empathy-Driven, Conversational Artificial Intelligence Agent (Wysa) for Digital Mental Well-Being: Real-World Data Evaluation Mixed-Methods Study.” JMIR MHealth and UHealth 6(11):e12106. doi:10.2196/12106.

Jang, Sooah, Jae-Jin Kim, Soo-Jeong Kim, Jieun Hong, Suji Kim, and Eunjoo Kim. 2021. “Mobile App-Based Chatbot to Deliver Cognitive Behavioral Therapy and Psychoeducation for Adults with Attention Deficit: A Development and Feasibility/Usability Study.” International Journal of Medical Informatics 150:104440. doi:10.1016/j.ijmedinf.2021.104440.

Johns Hopkins Medicine. 2025. “Kids Turning to Chatbot Therapy.” https://www.hopkinsmedicine.org/news/articles/2025/09/kids-turning-to-chatbot-therapy.

Koh, Jerica, Germaine Y. Q. Tng, and Andree Hartanto. 2022. “Potential and Pitfalls of Mobile Mental Health Apps in Traditional Treatment: An Umbrella Review.” Journal of Personalized Medicine 12(9). doi:10.3390/jpm12091376.

Lowrie, Karen, and Brooke Schwartzman. 2023. Youth Mental Health in New Jersey: Current Status and Opportunities for Improved Services. New Brunswick, NJ: Rutgers University. https://www.nj.gov/education/safety/wellness/mh/docs/NJSPL_YouthMentalHealth.pdf.

Lynch, Shana. 2025. Closing the Digital Divide in AI. https://hai.stanford.edu/news/closing-the-digital-divide-in-ai.

Marshall, Candace. 2025. “Chatbots vs. Conversational AI: What’s the Difference?” https://www.zendesk.com/blog/chatbot-vs-conversational-ai/.

Mennella, Ciro, Umberto Maniscalco, Giuseppe De Pietro, and Massimo Esposito. 2024. “Ethical and Regulatory Challenges of AI Technologies in Healthcare: A Narrative Review.” Heliyon 10(4):e26297. doi:10.1016/j.heliyon.2024.e26297.

National Institute of Mental Health. 2024. “Mental Illness.” https://www.nimh.nih.gov/health/statistics/mental-illness.

Newport Academy. 2024. “AI and Teen Mental Health: The Pros and Cons.” https://www.newportacademy.com/resources/empowering-teens/ai-teen-mental-health/.

De Nieva, Johan Oswin, Jose Andres Joaquin, Chaste Bernard Tan, Ruzel Khyvin Marc Te, and Ethel Ong. 2020. “Investigating Students’ Use of a Mental Health Chatbot to Alleviate Academic Stress.” Pp. 1–10 in 6th International ACM In-Cooperation HCI and UX Conference. New York, NY, USA: ACM.

Orsolini, Laura, Simone Pompili, Virginio Salvi, and Umberto Volpe. 2021. “A Systematic Review on TeleMental Health in Youth Mental Health: Focus on Anxiety, Depression and Obsessive-Compulsive Disorder.” Medicina (Kaunas, Lithuania) 57(8). doi:10.3390/medicina57080793.

Pava, Juan, Caroline Meinhardt, Haifa Badi Uz Zaman, Tori Friedman, Sang T. Truong, Daniel Zhang, Elena Cryst, Vukosi Marivate, and Sanmi Koyejo. 2022. Mind the (Language) Gap: Mapping the Challenges of LLM Development in Low-Resource Language Contexts. https://hai.stanford.edu/policy/mind-the-language-gap-mapping-the-challenges-of-llm-development-in-low-resource-language-contexts.

Potts, Courtney, Carmen Kealy, Jamie M. McNulty, Alba Madrid-Cagigal, Thomas Wilson, Maurice D. Mulvenna, Siobhan O’Neill, Gary Donohoe, and Margaret M. Barry. 2025. “Digital Mental Health Interventions for Young People Aged 16-25 Years: Scoping Review.” Journal of Medical Internet Research 27:e72892. doi:10.2196/72892.

Sanford, John. 2025. “Why AI Companions and Young People Can Make for a Dangerous Mix.” https://med.stanford.edu/news/insights/2025/08/ai-chatbots-kids-teens-artificial-intelligence.html.

Smith, Molly G., Thomas N. Bradbury, and Benjamin R. Karney. 2025. “Can Generative AI Chatbots Emulate Human Connection? A Relationship Science Perspective.” Perspectives on Psychological Science. doi:10.1177/17456916251351306.

Tam, Mallorie T., Julia M. Wu, Cindy C. Zhang, Colleen Pawliuk, and Julie M. Robillard. 2024. “A Systematic Review of the Impacts of Media Mental Health Awareness Campaigns on Young People.” Health Promotion Practice 25(5):907–20. doi:10.1177/15248399241232646.

